# Extra-axial inflammatory signal and its relation to peripheral and central immunity in depression

**DOI:** 10.1101/2024.03.15.24304342

**Authors:** Brandi Eiff, Edward T. Bullmore, Menna R. Clatworthy, Tim D. Fryer, Carmine M. Pariante, Valeria Mondelli, Lucia Maccioni, Nouchine Hadjikhani, Marco L. Loggia, Michael A. Moskowitz, Emiliano Bruner, Mattia Veronese, Federico E. Turkheimer, NIMA Consortium, Julia J. Schubert

## Abstract

Although both central and peripheral inflammation have been consistently observed in depression, the relationship between the two remains obscure. Extra-axial immune cells may play a role in mediating the connection between central and peripheral immunity. This study investigates the potential roles of calvarial bone marrow and parameningeal spaces in mediating interactions between central and peripheral immunity in depression.

Positron emission tomography was employed to measure regional TSPO expression in the skull and parameninges as a marker of inflammatory activity. This measure was correlated with brain TSPO expression and peripheral cytokine concentrations in a cohort enriched for heightened peripheral and central immunity comprising 51 individuals with depression and 25 healthy controls.

The findings reveal a complex relationship between regional skull TSPO expression and both peripheral and central immunity. Facial and parietal skull bone TSPO expression showed significant associations with both peripheral and central immunity. TSPO expression in the confluence of sinuses was also linked to both central and peripheral immune markers. Group dependant elevations in TSPO expression within the occipital skull bone marrow were also found to be significantly associated with central inflammation.

Significant associations between immune activity within the skull, parameninges, parenchyma, and periphery highlight the role of the skull bone marrow and venous sinuses as pivotal sites for peripheral and central immune interactions.

## Introduction

Ample evidence supports the theory that major depressive disorder (MDD) is, at least in part, linked to immune dysfunction.^1^ Heightened peripheral and central inflammation have been consistently observed among MDD cohorts.^1-12^ However, while significant elevations in peripheral and central inflammatory markers have been observed, they often do not appear to be directly correlated,^7-10,13^ and the link between the two phenomena remains unclear. Therefore, this work investigates whether an intermediate component is involved in the interplay between systemic and central inflammation in depression, and whether immune cells directly acting on the brain are sourced more locally from extra-axial regions.

Supporting the theory of an inflammatory aetiology of depression, numerous meta-analytic studies report elevated levels of peripheral pro-inflammatory cytokines in patients with depressive disorders.^2-6^ While previous evidence shows a dysregulation of nearly all immune markers within these cohorts,^2^ C-reactive protein (CRP), interleukin (IL)-6, and tumour necrosis factor (TNF)-α are among the most reliable inflammatory biomarkers adopted in depression studies.^1^ The production pathways of these acute-phase proteins are all interconnected, with increased concentrations of some regulating the release of others, ultimately resulting in systemic inflammation. TNF-α is a major pro-inflammatory cytokine produced by dendritic cells and macrophages and is involved in activating downstream inflammatory cascades, including the release of IL-6 from macrophages.^2,3^ IL-6, which can have pleiotropic functions pertaining to immune modulation, is produced by a myriad of cell types within the periphery and the brain.^14^ Following this specific lineage, the release of IL-6 from macrophages triggers the production of CRP by liver hepatocytes.^15^ Although clinical and immunopsychiatric research primarily utilizes CRP as a marker of inflammation, other markers may better capture the systemic immune dysfunction observed in depression.^16^

The relationship between depression and peripheral inflammation appears to have a bidirectional nature.^17^ The development of depressive symptoms is a prevalent side effect of immunotherapy, as observed when interferon (IFN)-α is administered to individuals being treated for non-psychiatric illnesses.^18,19^ Conversely, anti-inflammatory medications have demonstrated efficacy in reducing depressive symptoms and improving antidepressant treatment response in some cases,^20^ particularly among patients with elevated peripheral CRP values.^21,22^

Substantial evidence also points to a notable increase in central inflammation among individuals with MDD.^1,7-12^ Microglia, the primary regulators of brain immune activity under both physiological and pathological conditions, play a pivotal role.^23^ The 18-kDa outer mitochondrial translocator protein (TSPO) displays consistent upregulation in various cell types conducive to neuroinflammatory activity, including microglia, activated astrocytes, mast cells, macrophages, and leucocytes.^24-29^ Positron emission tomography (PET) studies utilizing radiotracers specific to TSPO have revealed evidence of neuroimmune activation across numerous neurological and psychiatric disorders.^30^ Notably, in MDD, increased TSPO expression has been reported within brain regions involved in mood regulation, including the anterior cingulate cortex (ACC), prefrontal cortex (PFC), and insula.^7,9^

Many researchers have investigated whether increased inflammatory activity in the brain is linked to peripheral immune responses.^31,32^ However, as reviewed by Turkheimer et al,^13^ much of the available data from human cohorts do not show a direct correlation between peripheral immune activation, as measured by plasma cytokines, and neuroinflammation, as measured by TSPO-PET. This observation has prompted a shift in focus towards understanding the functional role of extra-axial immune cells, originating from both the skull bone marrow and the meninges, in mediating the connection between systemic and central inflammation.

Such interest stems from compelling evidence from several recent preclinical studies that implicates immune cell trafficking across the skull and meninges in neuroinflammatory responses.^33-39^ Localized cell tagging of leukocytes has revealed that neutrophils recruited to inflamed cerebral tissue following stroke were primarily derived from skull bone marrow rather than distal tibial marrow.^35^ Additionally, neutrophils were shown to migrate from the skull to the brain, against typical blood flow, via direct vascular channels connecting skull bone marrow to the dura. Further investigation into this system has revealed that myeloid cell reservoirs exist within skull bone marrow which, under homeostatic conditions, supply monocytes and neutrophils to dural meninges.^34^ Moreover, during pathological states, meningeal niches dispense immune cells into brain parenchyma and are replenished by adjacent skull bone marrow, all seemingly independent of blood circulation or other peripheral immune responses. Importantly, communication along the skull-meninges-brain axis appears to be bidirectional, meaning that inflammatory signalling molecules from the brain carried through cerebrospinal fluid (CSF) also enter the skull, resulting in exacerbated immune responses from the skull bone marrow.^35,36,39^ Together, this evidence suggests that both the skull bone marrow and meninges play crucial roles in brain immunity, encompassing both homeostatic maintenance and responses to pathological conditions.

Pro-inflammatory cells entering the brain appear to be directly sourced from adjacent regions in the skull and meninges rather than the periphery. This hypothesis has been confirmed in a preclinical model of depression^37^ and could explain why previous studies have been unable to find significant direct correlations between central and peripheral inflammation. Hence, the link between skull, meningeal, and parenchymal immune compartments may provide a new perspective on inflammatory responses in MDD as well as novel therapeutic targets.

Extra-axial inflammatory responses during human neurological disease have recently been investigated using TSPO-PET. Disease-specific patterns of TSPO signals have been detected in the skull within stroke, multiple sclerosis, and neurodegenerative disease patients.^36^ In a study performed by Hadjikhani et al^40^ investigating the inflammatory correlates of migraine with visual aura, TSPO-PET signal was elevated within the meninges and skull bone marrow (parameningeal tissue [PMT]) overlying the occipital lobe. These findings, along with evidence of vasculature linking the skull and parenchyma, suggest an important interplay exists between upregulated inflammatory responses within the skull and the pathophysiology of diseases associated with neuroinflammation.^40^ This also may relate to previous findings of parameningeal immune mediating cell migration into the brain in a preclinical model of depression.^37^

These studies have provided a valuable framework for the present study to investigate whether these disease-associated patterns of TSPO expression in the calvaria bone marrow are echoed in a neuropsychiatric condition known to have an inflammatory component, such as depression. Hence, in this work, we investigated the association between the inflammatory status of parameningeal spaces and peripheral and central immunity in a large cohort of depressed subjects enriched for peripheral cytokines and neuroinflammatory activity. Correlations were then computed to examine the relationship between TSPO expression in the skull and parameningeal regions and both brain TSPO expression and peripheral cytokine concentrations.

## Methods and materials

### Participants

The study cohort comprised 51 depressed individuals (DP) aged 25 to 50 years, and 25 age-matched healthy controls (HC). Recruitment for this investigation was conducted across an array of clinical research sites in the United Kingdom, as part of the BIODEP (Biomarkers in Depression) study under the NIMA consortium (https://www.neuroimmunology.org.uk/biodep/). Depressed individuals recruited for this study exhibited a total Hamilton Depression Rating Scale (HDRS) score equal to or greater than 13. Depressed subjects representing a range of peripheral inflammatory states, as indicated by CRP levels, were enrolled. CRP concentration was adopted as a measure of the severity for the peripheral inflammatory state and depressive patients were categorized into high and low CRP groups using a predefined CRP concentration threshold of 3 mg/L. The HC participants were matched in terms of mean age to the DP and had no personal history of treated clinical depression.

Exclusion criteria for both the DP and HC participants included no documented history of neurological disorders, absence of current drug and/or alcohol abuse, no recent participation in clinical drug trials, absence of concurrent medication or medical conditions that could introduce confounding variables into result interpretation, and no current status of pregnancy or breastfeeding. Ethical endorsement for this study was secured from the National Research Ethics Service Committee East of England – Cambridge Central (REC reference: 15/EE/0092), and the research design adhered to the directives set forth by the UK Administration of Radioactive Substances Advisory Committee. Before data collection commenced, all participants granted written informed consent. Demographic and clinical characteristics are presented in detail in Table 1. Further insights into the dataset leveraged for this study can be referenced from the primary publication of the PK11195 PET results concerning this specific cohort.^9^

**Table 1.** Demographic, Clinical, and Scanning Characteristics.

| Variable<br>mean (SD) | Depressed<br>(*n* = 51) | Healthy<br>(*n* = 25) | *p* |
| --- | --- | --- | --- |
| **Demographic** |  |  |  |
| Age, years | 36.2 (7.3) | 37.3 (7.8) | 0.561 |
| Male, *n* (%) | 15 (29%) | 11 (44%) | 0.208 |
| Weight, kg | 80.3 (14.4) | 73.7 (15.1) | 0.072 |
| BMI, kg/m<sup>2</sup> | 27.7 (4.0) | 24.8 (3.9)<sup>a</sup> | 0.001 |
| **Clinical** |  |  |  |
| CRP, mg/L | 2.9 (2.8) | 1.1 (0.9) | < 0.001 |
| HDRS | 18.5 (3.7) | 0.6 (0.9) | <0.001 |
| State Anxiety Score | 50.7 (9.8) | 25.7 (6.2) | <0.001 |
| Trait Anxiety Score | 60.9 (7.9) | 28.9 (5.8) | <0.001 |
| **Scanning** |  |  |  |
| Dose, MBq | 360.3 (53.2) | 376.2 (44.8) | 0.138 |
| Injected Mass, mg | 3.0 (1.6) | 3.4 (1.8) | 0.486 |
| Specific Activity, GBq/mmol | 50.80 (21.33) | 50.56 (25.94) | 0.711 |
| Total Motion, mm | 7.76 (3.75) | 7.27 (3.39) | 0.562 |
| Max Interframe Motion, mm | 1.62 (0.90) | 1.45 (0.68) | 0.532 |
| Scan Start Time | 15:14:44 | 14:58:05 | 0.761 |

<sup>a</sup>BMI was not available for one control subject due to missing height measurement.
BMI = body mass index, CRP = C-reactive protein, HDRS = Hamilton depression rating score.

### Clinical assessments

Venous blood samples were collected from the antecubital region to measure levels of circulating peripheral inflammatory proteins and pro-inflammatory cytokines including CRP, TNF-α, and IL-6. Blood collection took place in the morning between 08:00 and 10:00 on the day of clinical assessment. To ensure standardized conditions, participants were instructed to fast for 8 hours and refrain from physical exercise for 72 hours before the blood sampling session. Subjects were required to lie in a supine position for 30 minutes before venous blood sample collection.

CRP levels were determined using a turbidimetry method conducted on Beckman Coulter AU analysers, employing latex particles coated with anti-CRP antibodies.^41^ For immune-related protein measurements, plasma preparation tubes (BD Cat #362799) were centrifuged at 1600g for 15 minutes at room temperature, and the resulting plasma supernatant was promptly frozen at -80°C. Upon thawing, markers were assayed in duplicate using the Pro-Inflammatory Panel 1 (K15049D) and Cytokine Panel 1 (K150150D) V-PLEX 10-spot immunoassay kits from Meso Scale Discovery, as per the manufacturer’s instructions (MSD; Rockville, MD, USA). Only samples with immune plasma concentrations equal to or exceeding the lower limit of detectability were included, which was 0.06 pg/mL for IL-6 and 0.05 pg/mL for TNF-α. The assay coefficients of variability were consistently below 15% for both biomarkers.^16^ Three healthy controls and one depressed subject were excluded due to unavailable IL-6 and TNF-α measurements.

### PET and MRI data acquisition

All participants underwent an imaging protocol consisting of a 60-minute simultaneous dynamic PET scan and high-resolution T1-weighted brain magnetic resonance imaging (MRI) scan following the administration of ^11^C-PK11195 (mean injected dose 361 ± 53 MBq) using a GE SIGNA PET/MR scanner (GE Healthcare, Waukesha, WI). For attenuation correction, a multi-subject atlas method was employed, incorporating enhancements for the MRI brain coil component.^42-44^ Concurrently, corrections for scatter, randoms, normalization, sensitivity, dead time, and decay were applied directly on the scanner. The dynamic sinograms were subsequently reconstructed into arrays with dimensions of 128 x 128 x 89, yielding a voxel size of 2 x 2 x 2 mm, utilizing the time-of-flight ordered subsets expectation maximization method with parameters set at 6 iterations, 16 subsets, and no smoothing.

### Image processing and quantification

Image pre-processing was performed using MIAKAT software (version 4.2.6; http://www.miakat.org/MIAKAT2/ index.html) within MATLAB version R2015b; The MathWorks, Inc., Natick, MA) and was completed as part of the original PET processing for the BIODEP study.^9^ Briefly, time activity curves were extracted from the bilateral ACC region using the CIC v2.0 neuroanatomical atlas transformed to subject image space. The ACC was chosen as the regional marker for brain inflammation based on previous results from this dataset.^9^ A more detailed account of image collection and initial processing can be found in the original report of the BIODEP PET data.^9^

Pseudo CT (pCT) images were synthesized from each subject’s T1 structural brain image using a fully automated online software (http://niftyweb.cs.ucl.ac.uk/).^42,43^ To isolate voxels primarily representing the skull, a predefined lower intensity threshold of 600 Hounsfield units was applied to the pCTs. The threshold was heuristically chosen based on voxel values in the skull and surrounding areas. Gaps or voids in the binarized image were filled to create a complete skull mask (Fig. 1).

**Figure 1.**
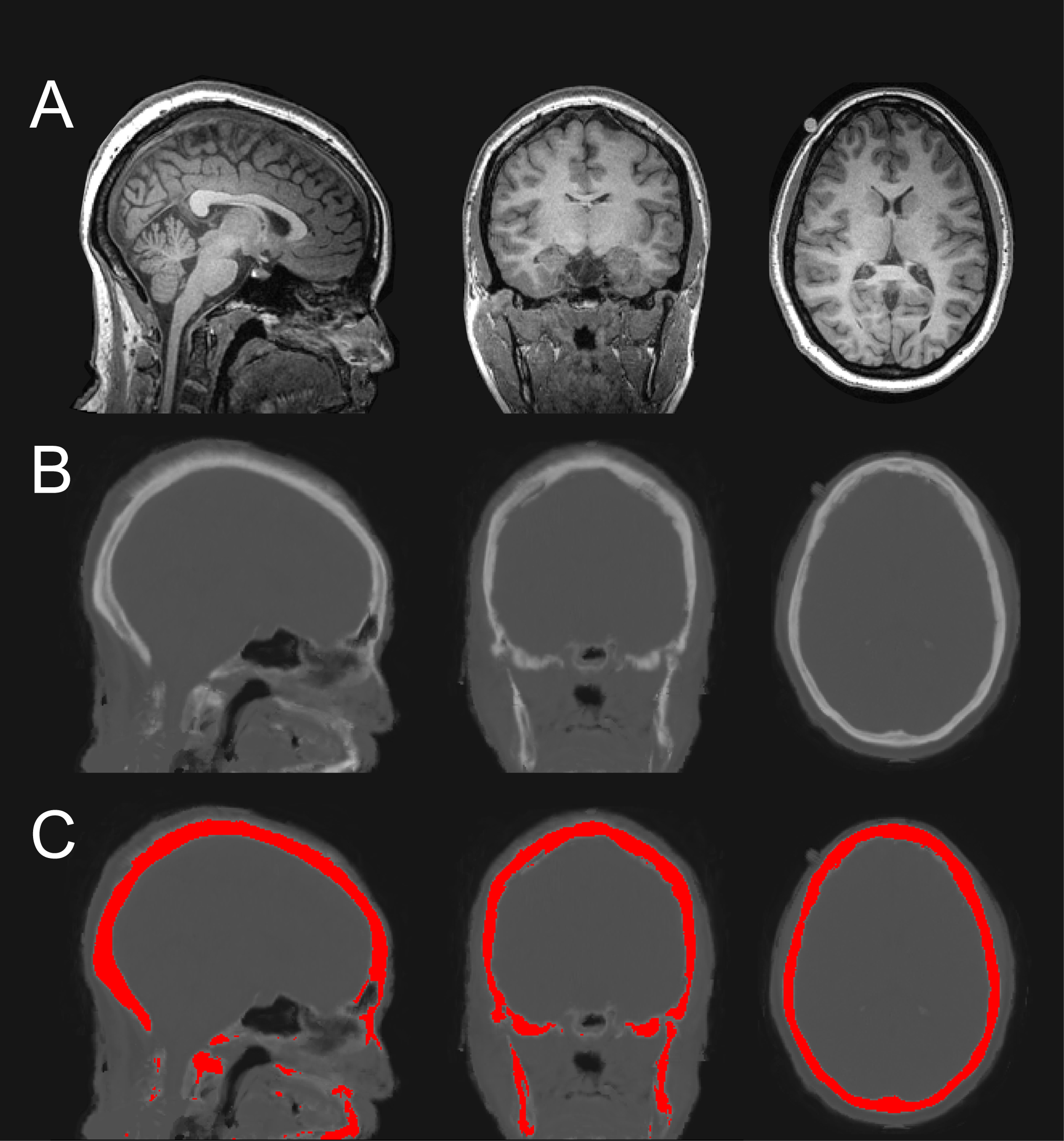
Generation of Skull Mask from Pseudo CT Images. (**A**) T1 structural brain image. (**B**) pCT image synthesized from T1 structural brain image. (**C**) pCT image with skull mask overlaid in red. Skull voxels were identified by applying a lower intensity threshold (600 Hounsfield units). Gaps and voids in the binarized image were filled to create a complete skull mask. pCt = pseudo computed tomography.

A PMT template devised as part of the previously discussed migraine study was provided by the HST/MGH A. A. Martinos Center for Biomedical Imaging.^40^ The ROIs included in this template consisted of the skull bone marrow and dura mater overlying the occipital lobe, parietal lobe, orbitofrontal cortex, and dorsolateral prefrontal cortex (DLPFC).

The PMT ROI templates were provided in 2-mm isotropic MNI space, and all ROIs originally had the same volume (100 voxels, 800 mm^3^). Additional processing of all ROIs was primarily performed using MATLAB version R2018b and associated toolboxes. Using SPM12 software, the templates were re-sliced to 1-mm isotropic resolution and non-linearly warped to align with the individual subjects’ structural MRI. These warped PMT templates were then dilated using a spherical kernel with radius of 3 voxels and masked using the previously defined skull ROI. This masking step was carried out to enhance the accuracy of the ROIs in depicting the regions of the skull. The ROIs were then eroded using a spherical kernel with radius of 1 voxel to mitigate the effects of partial volume of surrounding tissues. The warped and masked PMT templates were then re-sliced again to 2-mm isotropic resolution to align with the subjects’ dynamic PET. It should be noted that the ROI signal analysed in this study is associated with gross regions of the bone covering the DLPFC, parietal, and occipital cortex. These bone regions have no established homological value or position and must be hence intended as a general sample of the frontal, parietal, and occipital squamae, respectively. In this sense, we use the terms “DLPFC skull”, “parietal skull” and “occipital skull” not to refer to the whole bones, but just to a minor central part of the overlying bones sampled in this study.

Segmentation was performed using FreeSurfer v6.0 software, utilizing as input the T1 structural brain images that had previously been co-registered to the subjects’ PET native space. A binary mask delineating non-brain areas was derived from FreeSurfer output by subtracting a binarized mask, obtained from the FreeSurfer parcellation image (aparc.a2009s+aseg_bin.nii.gz), from the FreeSurfer brainmask image.

This mask, initially encompassing all non-brain spaces, was further refined to specifically isolate the posterior cranial fossa. This region houses the confluence of sinuses, which is recognized as the largest venous blood pool, enabling the quantification of TSPO expression while mitigating the effects of partial volume. Clusters smaller than 5 voxels within the posterior non-brain mask were excluded, and the retained clusters were further dilated with a spherical kernel having a radius of 4 voxels to ensure full coverage of the confluence of sinuses. Voxels were considered to belong to the same cluster if they shared at least one edge. Dynamic PET data were subsequently utilized to identify and preserve voxels within the mask that exhibited activity of at least 25% of the peak activity observed in the frame with the highest number of voxels displaying maximum activity (the mode peak frame time). The kinetics of these voxels strongly suggest that they predominantly contain blood. Smaller clusters containing fewer than 5 voxels, attributed to noise, were removed from the mask. To refine the mask’s edges, a process was applied involving dilation followed by erosion using a spherical kernel with a radius of 2 voxels. Finally, only the largest volume cluster was retained in the mask resulting in the final binary mask representing the confluence of sinuses ROI (Fig. 2).

**Figure 2.**
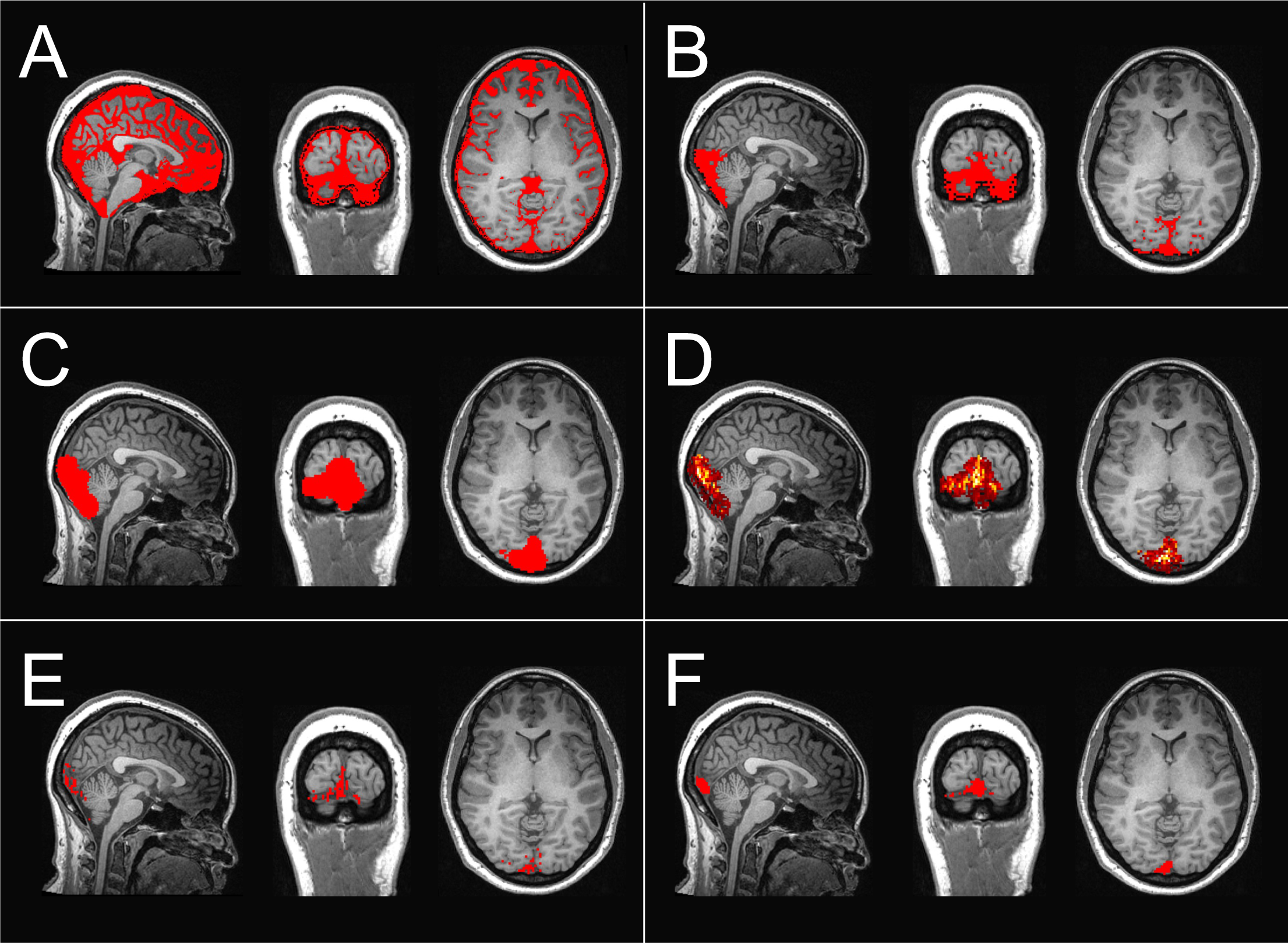
Confluence of Sinuses ROI Generation Process. (**A**) Binary mask delineating non-brain areas, overlaid on T1 structural brain image. (**B**) Refinement of the mask to isolate the posterior cranial fossa. (**C**) Exclusion of small clusters and dilation of retained clusters to ensure coverage of the confluence of sinuses. (**D**) Employing PET data to identify voxels exhibiting at least 25% of the peak activity observed in the mode peak frame time, indicating a predominant presence of blood in those voxels. (**E**) Only voxels identified as predominantly containing blood are retained in binary mask. (**F**) Removal of smaller noise clusters, refinement of mask edges through dilation and erosion, and retention of the largest volume cluster to produce the final binary mask representing the confluence of sinuses. PET = positron emission tomography, ROI = region of interest.

A facial bone ROI was defined by excluding voxels associated with the neurocranium while specifically including bones belonging to the upper viscerocranium. This resulted in an ROI comprising the glabella, nasal bones, zygomatic bones, lacrimal bones, coronoid processes of the mandible, ethmoid bone, sphenoid bone, vomer, and the maxillary body. All ROIs (Fig. 3) underwent thorough visual inspection for quality assurance, and manual adjustments were made as necessary to ensure the accurate representation of intended tissues within the ROIs.

**Figure 3.**
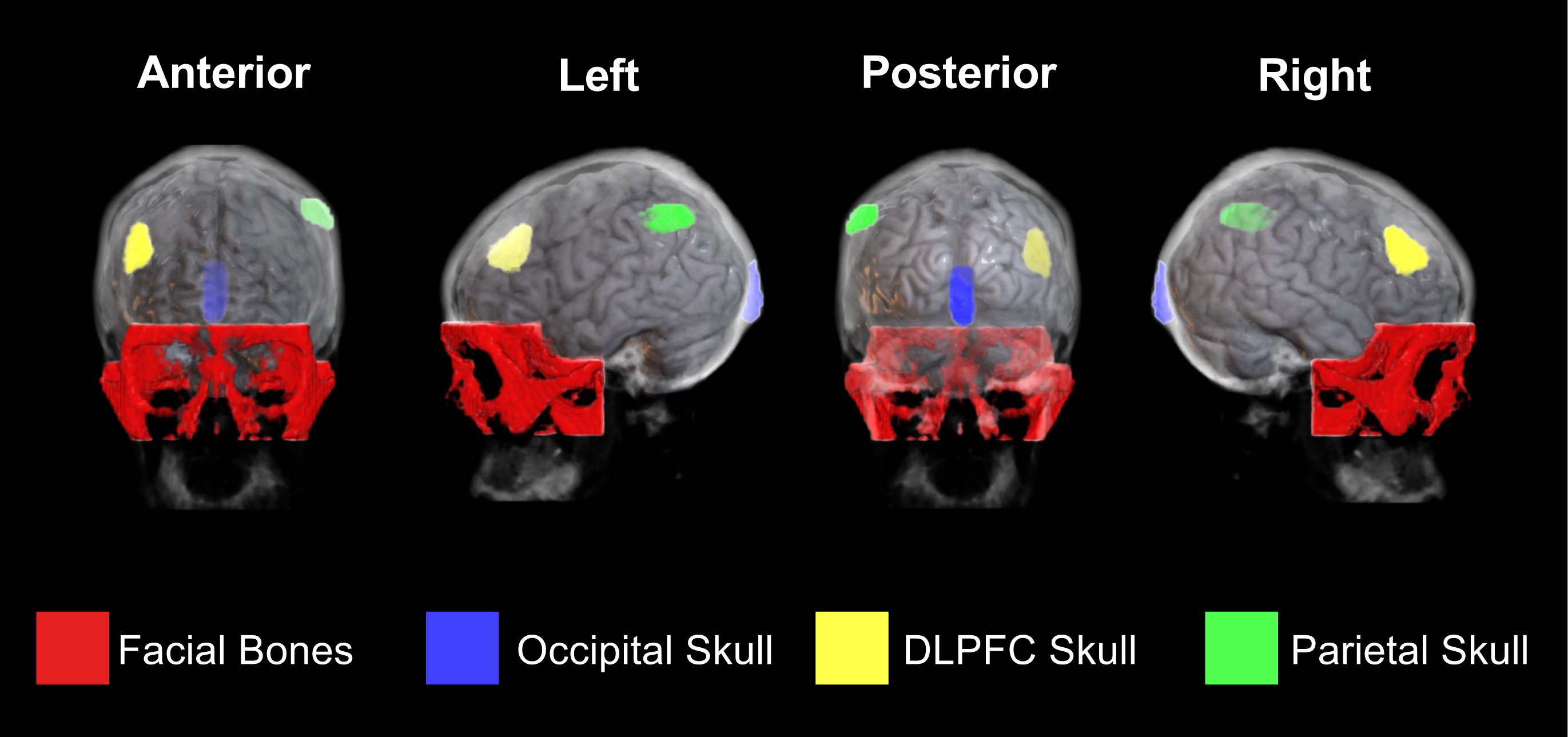
3D Representation of Skull Regions of Interest. Three-dimensional rendering depicting specific regions of interest within the skull. Regions of interest were informed by template provided by Hadjikhani et al ^40^ and pseudo CT renderings derived from participants’ structural MRIs. The use of pseudo CTs facilitated the refinement of these regions, ensuring that voxels exclusively represented bone marrow without including adjacent dura mater. The confluence of sinuses was identified by selecting voxels with PET kinetics indicating a predominant presence of blood within the posterior cranial fossa. DLPFC = dorsolateral prefrontal cortex, MRI = magnetic resonance imaging.

Time activity curves (TACs) were extracted for each subject across occipital, DLPFC, and parietal skull regions, facial bone, and confluence of sinuses ROIs as well as the ACC. Quantification of ^11^C-PK11195 binding within these regions was accomplished using standardized uptake values (SUVs), using TAC data from 5 minutes to the end of the scan time in order to mitigate the impact of early frame noise and tracer delivery. Note that in previous studies intra-axial activity was normalized to a brain reference region; in this work this approach was avoided to ensure consistency in the quantification method for both extra- and intra-axial regions.

### Statistical analysis

All statistical analyses were carried out using SPSS software (version 28.0.1.1; IBM, Armonk, NY). The normality of all dependent variables was assessed using the Shapiro-Wilk test. Group differences in trait anxiety and body weight were analysed using the independent-samples *t*-test. All other group differences in experimental variables, encompassing demographic, clinical, and scanning characteristics, were evaluated using the independent-samples Mann-Whitney U test (Table 1). An extreme outlier with a notably high TNF-α value was identified in one DP case and excluded from further analysis.

A repeated measures general linear model (GLM) was employed, treating extra-axial regional average SUV values (facial bones, occipital skull, DLPFC skull, parietal skull, and confluence of sinuses) as the repeated measure, subject group as the fixed factor, and TNF-α, IL-6, CRP, and ACC SUV as covariates. Before being included in statistical analyses, CRP and cytokine values underwent log10 transformation to achieve an approximately normal distribution. Only variables that exhibited significant contributions (*p*-value ≤0.05) to the statistical model were retained in the final model.

Additionally, a univariate analysis of covariance was conducted for the average SUV in each region (facial bones, occipital skull, DLPFC skull, parietal skull, and confluence of sinuses), with subject’s group as the fixed factor and TNF-α and ACC SUV as covariates. A Spearman’s correlation was conducted to investigate the relationship between TNF-α and ACC SUV. All p-values are reported without correction for multiple comparisons.

## Results

A repeated measures GLM was performed to determine whether there were statistically significant group differences in TSPO signal across extra-axial regions and whether central and peripheral inflammatory markers contributed to these variations. Initially, this test was performed including CRP, IL-6, TNF-α, and ACC SUV as covariates, with the group as a between-subjects factor. However, tests of between-subjects effects revealed that CRP and IL-6 did not have a significant main effect on TSPO signal variability across regions [CRP: *F*(1, 64) = 0.981, *p* = 0.326, η*p*² = .015; IL-6: *F*(1, 64) = 1.572, *p* = 0.215, η*p*² = .024] and were therefore excluded from the final model.

The GLM was performed again including TNF-α and ACC SUV as covariates and group as a between-subjects factor. Mauchly’s test of sphericity indicated that the assumption of sphericity was violated [χ*^2^*(9) = 84.928, *p* <0.001] and therefore Greenhouse-Geisser correction was applied (ε = 0.657). Results from the GLM indicated that TNF-α, ACC SUV, and group significantly contributed to variation in extra-axial inflammation between subjects [TNF-α: *F*(1, 67) = 4.747, *p* = 0.033, η*p*² = 0.066; ACC SUV: *F*(1, 67) = 14.263, *p* < 0.001, η*p*² = 0.176; group: *F*(1, 67) = 4.005, *p* = 0.049, η*p*² = 0.056]. A significant interaction effect was also observed between the extra-axial region and ACC SUV [*F*(2.628, 176.083) = 3.312, *p* = 0.027, η*p*² = 0.047], prompting additional univariate analyses within each specific extra-axial region.

Univariate analysis of covariance (ANCOVA) was used to assess whether inflammatory signal within the individual extra-axial ROIs was significantly influenced by peripheral and central inflammation, represented by TNF-α and ACC SUV respectively, and group. ACC SUV had a significant positive effect on SUV values within the occipital skull [*F*(1,67) = 4.667, *p* = 0.034, η*p*² = 0.065], facial bones [*F*(1,67) = 7.546, *p* = 0.008, η*p*² = 0.101], parietal skull [*F*(1,67) = 6.504, *p* = 0.013, η*p*² = 0.088], and the confluence of sinuses [*F*(1,67) = 118.883, *p* = < 0.001, η*p*² = 0.640] (Fig. 4). These values indicate that TSPO signal within the ACC is significantly correlated with TSPO signal in these regions, irrespective of group differences, with the most robust association found in the confluence of sinus region (Table 2).

**Figure 4.**
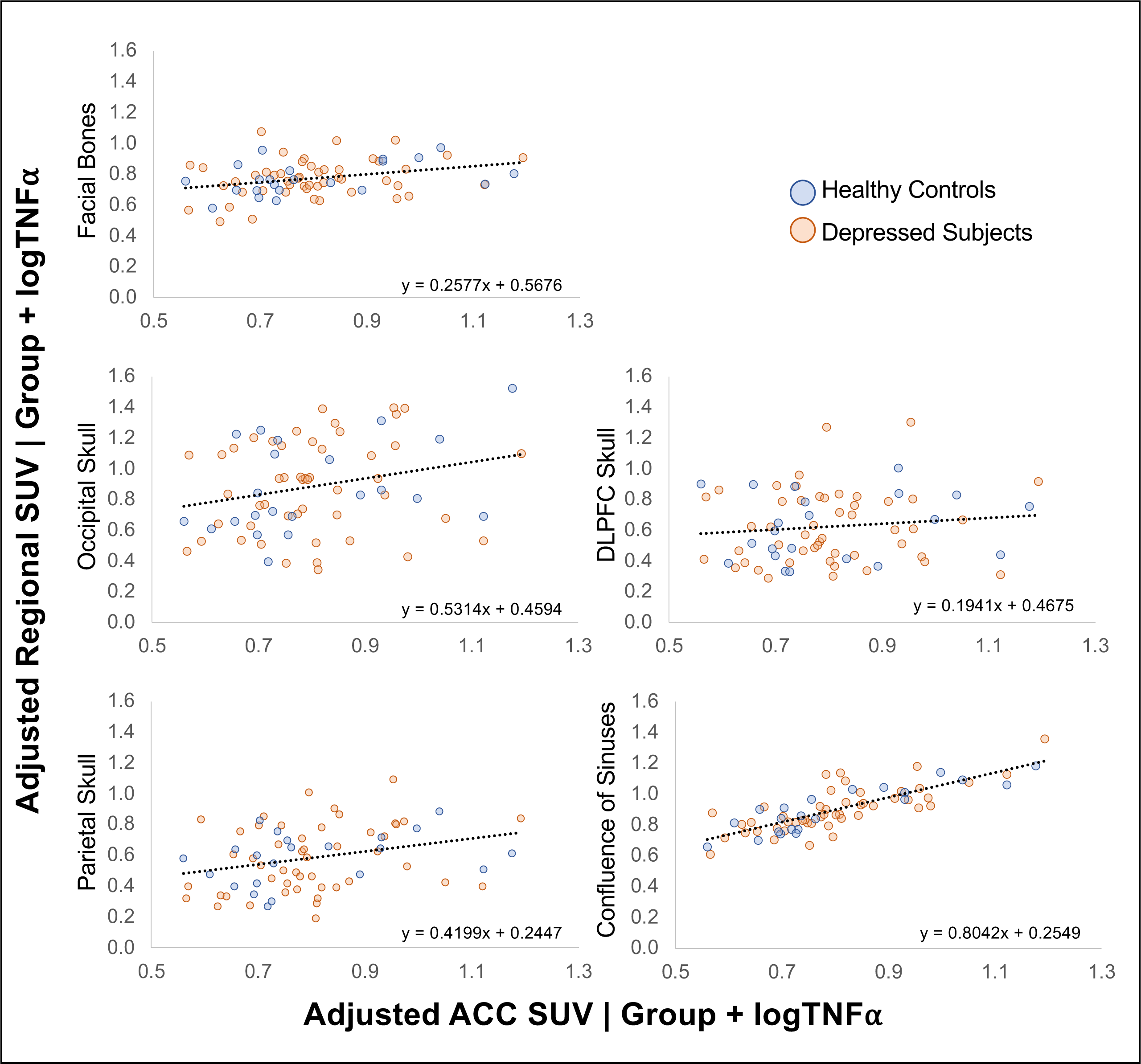
ACC SUV Effect on Regional Extra-Axial SUV. Partial regression plots presenting the relationship between ACC SUV and regional extra-axial SUV, accounting for group and logTNFα. ACC = anterior cingulate cortex, SUV = standardized uptake value.

**Table 2.**
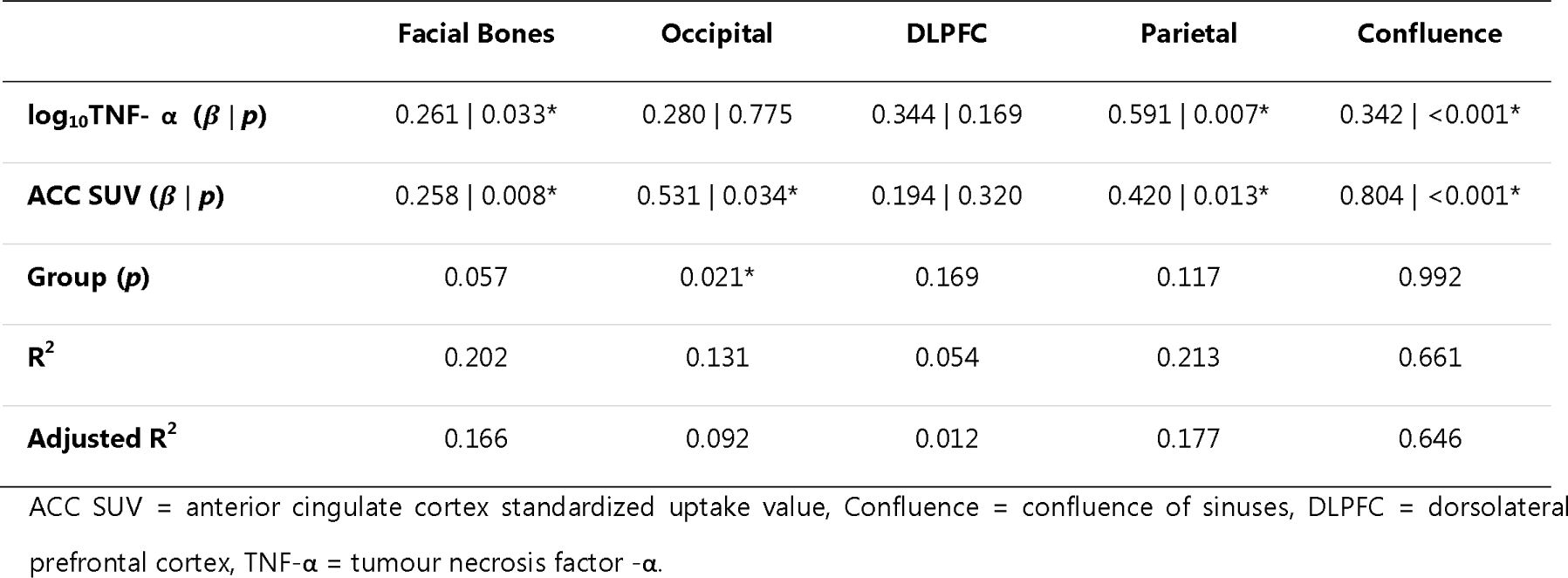
Overview of univariate results.

|  | Facial Bones | Occipital | DLPFC | Parietal | Confluence |
| --- | --- | --- | --- | --- | --- |
| **log<sub>10</sub>TNF- α (β \| *p*)** | 0.261 \| 0.033* | 0.280 \| 0.775 | 0.344 \| 0.169 | 0.591 \| 0.007* | 0.342 \| <0.001* |
| **ACC SUV (β \| *p*)** | 0.258 \| 0.008* | 0.531 \| 0.034* | 0.194 \| 0.320 | 0.420 \| 0.013* | 0.804 \| <0.001* |
| **Group (*p*)** | 0.057 | 0.021* | 0.169 | 0.117 | 0.992 |
| **R<sup>2</sup>** | 0.202 | 0.131 | 0.054 | 0.213 | 0.661 |
| **Adjusted R<sup>2</sup>** | 0.166 | 0.092 | 0.012 | 0.177 | 0.646 |

ACC SUV = anterior cingulate cortex standardized uptake value, Confluence = confluence of sinuses, DLPFC = dorsolateral prefrontal cortex, TNF-α = tumour necrosis factor -α.

TNF-α also has a significant positive effect on SUV variance within the facial bones [*F*(1,67) = 4.751, *p* = 0.033, η*p*² = 0.066], parietal skull [*F*(1,67) = 7.876, *p* = 0.007, η*p*² = 0.105], and confluence of sinuses [*F*(1,67) = 13.142, *p* < 0.001, η*p*² = 0.164] (Fig. 5). No significant association was found between TNF-α and ACC SUV [*r*(71) = -0.072, *p* = 0.553].

**Figure 5.**
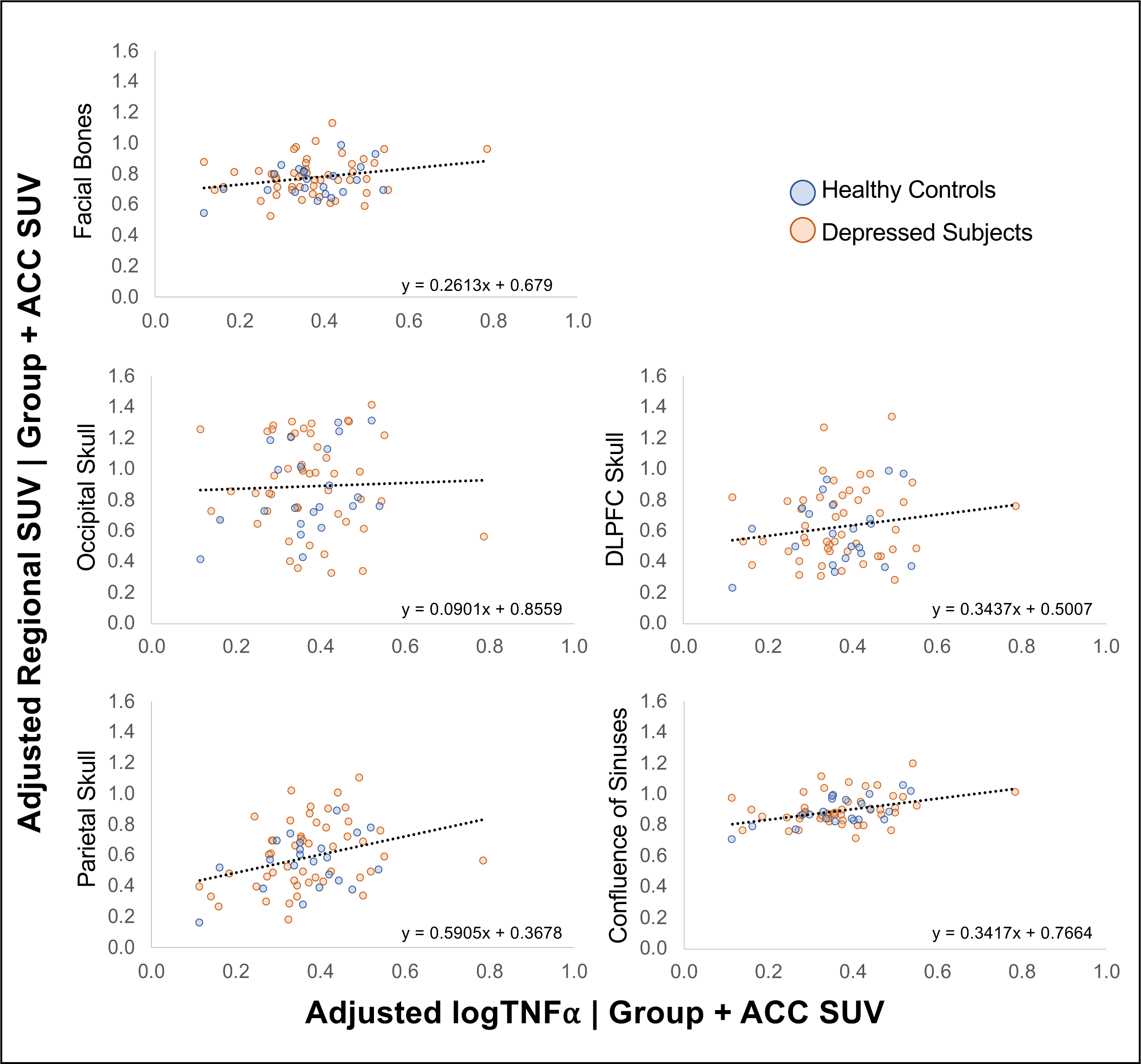
logTNFα Effect on Regional Extra-Axial SUV. Partial regression plots presenting the relationship between logTNFα and regional extra-axial SUV, accounting for group and ACC SUV. ACC = anterior cingulate cortex, SUV = standardized uptake value.

The univariate ANCOVA revealed a significant main effect of group on occipital skull SUV only [*F*(1, 67) = 5.605, *p* = 0.021, η*p*² = 0.077]. Participants in the depression group exhibited higher occipital skull SUV values (*M* = 0.975, *SE* = 0.043) than controls (*M* = 0.791, *SE* = 0.064), after adjusting for ACC SUV and TNF-α (Fig. 6).

**Figure 6.**
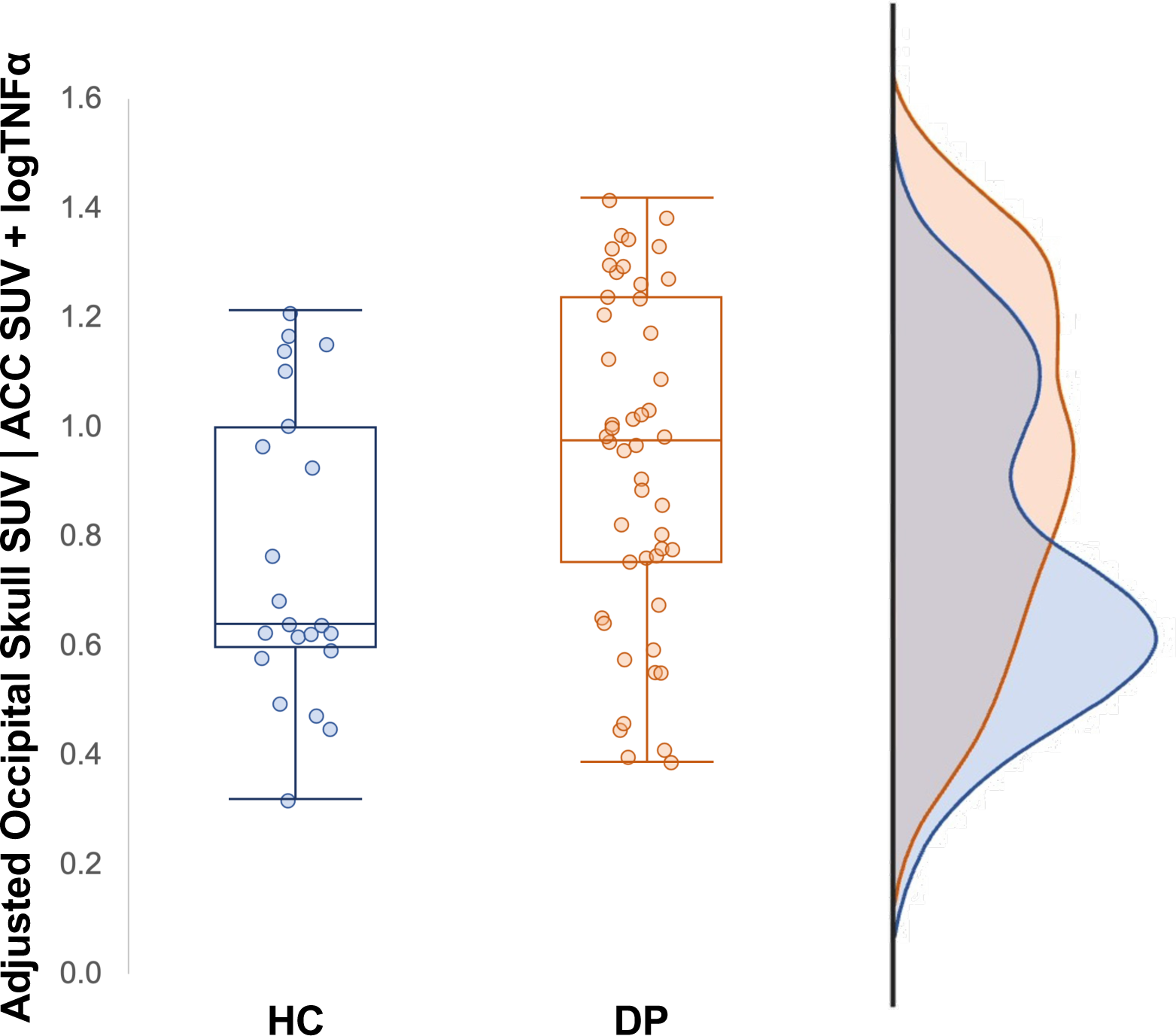
Group Effect on Occipital Skull SUV. Group differences in occipital skull SUV, adjusted for ACC SUV and logTNFα. ACC = anterior cingulate cortex, DP = depressed individuals, HC = healthy controls, SUV = standardized uptake value.

## Discussion

The present study examined the relationship between skull bone marrow immune activity and peripheral as well as central inflammation within a cohort of depressed individuals and healthy controls. The dataset included a diverse range of peripheral and central inflammatory states, as evidenced by CRP levels and parenchymal TSPO expression, respectively, providing valuable insights into the interplay between these factors. The results suggest that calvaria marrow, the source of TSPO signal in the skull, and venous blood pool are associated with both peripheral and central immune signals. However, this association varies regionally.

A robust significant association was observed between the TSPO-PET signal within the confluence of sinuses and the parenchymal ACC inflammatory signal, as well as with peripheral TNF-α concentrations. Located along the occipital bone, the confluence of sinuses plays a vital role in draining endocranial blood, generally through the lateral or occipito-marginal sinuses, often both converging into the jugular canals.^45-47^ Accordingly, whatever physiological product introduced into this sinus is quickly drained out of the braincase. Thus, it is reasonable to infer that the signal detected may be associated with the perivascular space of the sinus, and not just with the sinus itself. The dural sinuses appear to be critically involved in immune cell surveillance, recruitment, and trafficking, acting as an interface facilitating interactions between the central and peripheral immune systems.^48^ The dural lymphatics likely also contribute to this connection, as they have previously been observed to form an extensive network at the base of the skull proximal to the dural venous network.^49^ Notably, ACC SUV and TNF-α measures, representing central and peripheral immune activity respectively, did not exhibit a direct correlation with each other. This finding aligns with previous reports that have struggled to establish a direct link between central and peripheral immunity.^9,13,50^ However, both ACC SUV and TNF-α showed strong positive associations with TSPO signal in the confluence of sinuses. These observations underscore the venous blood pool as an important hub for central-peripheral immune crosstalk.

We observed weaker associations with peripheral and central immunity in the marrow of facial bones and the parietal skull. The recently uncovered network of microscopic vascular channels, documented in frontal, parietal, and occipital skull regions, serves as a conduit for communication between these systems.^33,35,38^ Regional variations in channel length, width, and density^39,51^ may account for our disparate findings across skull regions. Our finding in parietal skull and facial bones suggests a subtle intermediary communication between central and peripheral systems. Notably, this study marks the first exploration of the potential involvement of facial bones in this context. The investigation was prompted by the proximity of facial bones to the nasal passages and sinuses, housing the nasal microbiota crucial for immune regulation and general CNS homeostasis.^52,53^ The TSPO signal in facial bones may be sensitive to changes in these spaces; however, further research is needed to elucidate the connection between facial bones and nasal passages and its implications for the brain-periphery connection.

A significant positive association with parenchymal ACC TSPO was found in the occipital skull region. A large hematopoietic niche containing a variety of immune cells has been observed within the caudal region of the skull in mice models^38^ which, in humans, translates to the occipital region. Moreover, in rodents the occipital skull region also contains a high density of skull channels that allow contact between the calvaria marrow and the CSF that is exiting the parenchyma and, during pathological states, enable immune messengers from the CNS to communicate with the periphery.^39^

TSPO expression in the frontal skull was not found to be correlated with TSPO levels in the underlying tissues of the frontal cortex. This observation may seem unexpected considering prior indications of similar TSPO patterns in skull and adjacent brain regions patients with Alzheimer’s disease.^36^ However, CSF serves as a dynamic communicator, playing a vital role not only in the immediate proximity but also in facilitating communication throughout the intricate network of the brain and its cranial environment. This finding also offers reassurance that the signals detected in the skull are not solely a consequence of partial volume effects of neighbouring tissues - a limitation associated with the PET scanner resolution.

Finally, we noted that depression group, as a statistical factor, was also a significant contributor to the increase in inflammation, specifically in the occipital skull and not in other extra-axial regions. This suggests that disease may modulate the local inflammatory response of the occipital skull through mechanisms beyond those considered in this study, involving factors beyond peripheral cytokines and neuroinflammation. Alternatively, the measurements of TNF-α concentrations and parenchymal TSPO expression may not provide a comprehensive representation of peripheral or central immune activity associated with depression.

The mechanism that has been hypothesized here may be transdiagnostic. For example, occipital PMT TSPO elevation has also been observed in patients with migraine with aura^40^ and may be the source of comorbidity across conditions (e.g. patients with depression have two-fold chance of developing migraine^54^ and there is a ±45% heritable association of migraine and MDD).^55^ Distinct patterns of TSPO signal elevation in the calvaria have also been observed in various pathologies, including multiple sclerosis, stroke, 4-repeat tauopathies, and Alzheimer’s disease.^36^ In instances of central inflammation, a phenomenon observed across these pathologies, there is cross talk between brain immune cells and immune cell reservoirs within the skull bone marrow, likely contributing to the elevated inflammatory signal that was previously observed in this depressed cohort.^9^

## Limitations

A few limitations should be considered in this study. Firstly, TSPO is expressed in numerous cell types and there is considerable debate as to the validity of its use as a conclusive marker for microglial activation. Other cell types which commonly express TSPO include astrocytes, activated macrophages, mast cells, neutrophiles, epithelial cells, and vascular endothelial cells.^27-29^ However, it is important to note that these cell types are all implicated in central immune mechanisms. Therefore, although TSPO expression may not solely represent microglial activation, it is reasonable to conclude that elevations in TSPO expression observed in this study are conducive to inflammatory activity. Furthermore, given that this study examined the skull bone marrow, this inherently limits the possible TSPO-expressing cell types. Therefore, this strengthens our confidence that the TSPO signal expressed in the skull marrow is representative of immune activity.

Another possible limitation in this study lies in the use of the radiotracer ^11^C-PK11195. Although second-generation TSPO tracers, such as ^11^C-PBR28 and ^18^F-DPA-714, have been shown to have higher sensitivity to TSPO,^30^ they present challenges when used in humans. Approximately one-third of human subjects carry genetic polymorphisms within the TSPO gene, leading to low binding potential in second-generation tracers.^56^ ^11^C-PK11195 circumvents these challenges, enabling easier between-subjects comparisons of TSPO binding. Importantly, numerous previous studies have validated the efficacy of ^11^C-PK11195 in measuring TSPO expression,^8^ supporting its utility in our investigation. The detection of significant group differences in our cohort using a less sensitive first-generation tracer adds robustness to our findings.

## Conclusion

In conclusion, this study’s findings provide in-vivo confirmation of observations from preclinical models, emphasising the role of calvaria marrow in facilitating brain-periphery immune interactions within a human cohort. Furthermore, the occipital region may harbour a special immune cell niche that is reserved for supplying immune privilege to the brain during disease states, consistent with previous findings. These results also underscore the significance of the parietal and facial bone marrow as well as the venous sinuses as key locations for this crosstalk, thereby highlighting these regions as potential targets for future treatment strategies in central immune-related conditions.

## Supporting information

Figure 3 - 3D Model

## Acknowledgements

We sincerely thank all who were involved in this study, including the participants, research teams, and laboratory staff, as their contributions were indispensable to the success of our research.

## Data availability

The data that support the findings of this study are available from the corresponding authors upon reasonable request.

