## Supplementary figures and images for "Extra-axial inflammatory signal and its relation to peripheral and central immunity in depression"

### Figure 3 - 3D Model

#### Slide 1
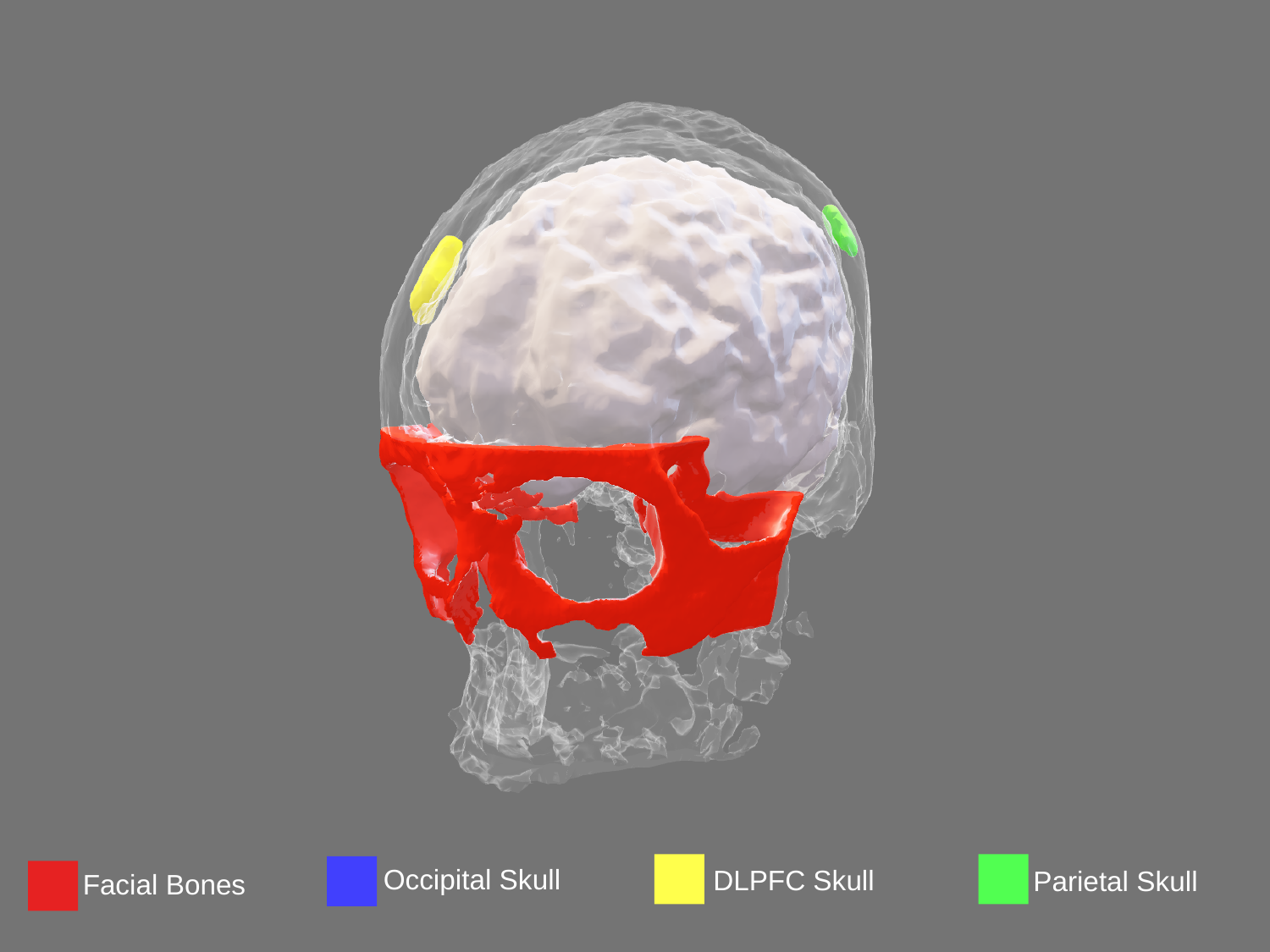

Occipital Skull
DLPFC Skull
Parietal Skull
Facial Bones
